# The molecular landscape of premature aging diseases defined by multilayer network exploration

**DOI:** 10.1101/2023.12.19.23300210

**Authors:** Cécile Beust, Alberto Valdeolivas, Anthony Baptista, Galadriel Brière, Nicolas Lévy, Ozan Ozisik, Anaïs Baudot

## Abstract

Premature Aging (PA) diseases are rare genetic disorders that mimic some aspects of physiological aging at an early age. Various causative genes of PA diseases have been identified in recent years, providing insights into some dysfunctional cellular processes. However, the identification of PA genes also revealed significant genetic heterogeneity and highlighted the gaps in our understanding of PA-associated molecular mechanisms. Furthermore, many patients remain undiagnosed. Overall, the current lack of knowledge about PA diseases hinders the development of effective diagnosis and therapies and poses significant challenges to improving patient care.

Here, we present a network-based approach to systematically unravel the cellular functions disrupted in PA diseases. Leveraging a network community identification algorithm, we delved into a vast multilayer network of biological interactions to extract the communities of 67 PA diseases from their 132 associated genes. We found that these communities can be grouped into six distinct clusters, each reflecting specific cellular functions: DNA repair, cell cycle, transcription regulation, inflammation, cell communication, and vesicle-mediated transport. We propose that these clusters collectively represent the landscape of the molecular mechanisms that are perturbed in PA diseases, providing a framework for better understanding their pathogenesis. Intriguingly, most clusters also exhibited a significant enrichment in genes associated with physiological aging, suggesting a potential overlap between the molecular underpinnings of PA diseases and natural aging.

## 1 Introduction

Premature Aging (PA) diseases are rare genetic disorders that exhibit clinical features mimicking physiological aging but manifesting at an early age[1,2]. The clinical resemblances observed between premature and physiological aging have attracted a lot of interest, with the idea that premature aging diseases may provide insights into the molecular features involved in physiological aging[1,3,2]. Aging phenotypes observed in PA diseases include, for instance, alopecia, hair graying, lipodystrophy, osteoporosis or atherosclerosis[3,2]. However, overall, PA syndromes are clinically heterogeneous, presenting diverse sets of phenotypes, affecting different tissues and organs, and exhibiting variations in the age of onset, progression and severity[4].

In recent years, advances in diagnosis techniques have allowed deciphering the genetic causes of various PA diseases. To date, over a hundred causative genes have been unveiled, revealing significant genetic heterogeneity[3]. Recent reviews have classified PA diseases in three major groups based on the molecular mechanisms associated with their causative genes[5,6]. The first group mainly comprises diseases associated with alterations of the nuclear lamina architecture. This first group includes syndromes associated with defective Prelamin A processing, such as Hutchinson-Gilford Progeria Syndrome (HGPS) and Restrictive Dermopathy, and other laminopathies such as Mandibuloacral Dysplasia or Néstor-Guillermo Progeria Syndrome. The second group encompasses diseases resulting from alterations of DNA repair pathways, such as Werner syndrome, Cockayne syndrome, Xeroderma Pigmentosum or Bloom syndrome. Finally, the third group gathers PA diseases associated with diverse molecular defects, including connective tissue alterations (Ehlers-Danlos syndromes and Cutis Laxa), impairments in mitochondrial pathways (Fontaine progeroid syndrome), or others. For some diseases in this last group, the mechanisms underlying a premature senescence are not fully elucidated[2]. Importantly, the genetic underpinnings of many PA diseases are still unknown, leaving numerous patients without a definitive molecular diagnosis[6,2]. Overall, gaining a deeper insight into the molecular mechanisms associated with PA diseases is crucial for enhancing diagnosis, optimizing patient care, and developing new therapeutic strategies to address the significant unmet needs in these ultra-rare genetic disorders.

The study of disease genes and proteins in their cellular network contexts provides a promising approach to understand the molecular mechanisms perturbed in diseases[7,8]. Indeed, proteins do not act as isolated molecules in cells but, instead, interact with each other to perform their cellular functions. Interactions are typically depicted as networks, in which the nodes represent individual genes or proteins, and the edges represent their physical or functional interactions. With the advancement of experimental techniques, biological interactions data accumulate and allow the construction of diverse biological interaction networks. These networks can be organised in multilayer networks that gather, for instance, protein-protein interactions, interactions occurring in molecular complexes, pathways, or gene/protein relationships inferred from –omics data, such as co-expression relationships inferred from transcriptomics data. This wealth of interaction data enables comprehensive studies of gene and protein cellular functions. However, given the magnitude and complexity of these interaction networks, appropriate computational methods are crucial for meaningful exploration.

Community detection is a broadly-used network analysis strategy. Biological networks tend to be structured into communities, which are subnetworks composed of nodes that share more connections with one another than with the rest of the network[9]. Communities in networks are expected to contain genes or proteins that are likely involved in the same processes or functions within cells. Hence, identifying these communities enables the exploration of cellular functions using the ‘guilt-by-association’ principle. Importantly, by pinpointing the communities encompassing disease-related genes, we gain a deeper insight into the cellular functions that are disrupted in these diseases[10]. A wide variety of computational methods have been proposed to extract communities from biological networks[11]. Recently, various community identification algorithms have been extended to consider as input multiple biological networks, thereby providing improved mapping of cellular functions[12,11,13,14].

Our aim here is to systematically unveil the cellular functions underlying PA diseases. To this end, we conducted a comprehensive analysis of 67 PA diseases and their 132 associated genes thanks to a network medicine strategy. Leveraging a novel community identification algorithm, we delved into a vast multilayer network of biological interactions, uncovering the 67 communities associated with the PA diseases. We next clustered the 67 disease communities based on their shared gene/protein nodes. This revealed a striking classification of the communities into six distinct clusters reflecting various cellular functions: DNA repair, cell cycle, transcription regulation, inflammation, cell communication, and vesicle-mediated transport. We propose that these clusters collectively represent the landscape of the dysfunctional molecular mechanisms in PA diseases. Our study thereby provides a valuable framework for decoding the pathogenesis of these disorders and guiding advancements in diagnostic and therapeutic approaches. Additionally, these clusters also exhibited a significant enrichment in genes associated with physiological aging, suggesting a potential overlap between the molecular underpinnings of PA diseases and the natural aging process.

## 2 Materials and methods

### 2.1 Premature aging disease and gene datasets

We inspected the Human Phenotype Ontology (HPO)[15] to collect phenotypes associated with Premature Aging (PA). We selected the HPO annotation term: “HP:0007495: prematurely aged appearance” together with all its descendant terms in the HPO ontology graph (Supplementary Table S1). In ORPHANET[16], we identified 87 diseases associated with at least one of these HPO terms. We called and hereafter refer to these 87 diseases as Premature Aging (PA) diseases. Focusing on the PA diseases having at least one associated gene in ORPHANET, we obtained 67 PA diseases associated with 132 genes (Supplementary Table S2). Using the 132 PA genes as input and all annotated genes as background, we performed functional enrichments with the g:Profiler web server (version `e110_eg57_p18_4b54a898`). Functional enrichments were computed for Gene Ontology[17] Biological Processes, Gene Ontology Cellular Components, and Reactome[18] annotations (Supplementary File S1). P-values were corrected using the Benjamini-Hochberg procedure.

### 2.2 Multilayer biological network

The diverse physical and functional interactions between genes and proteins can be represented as a collection of network layers[8,19]. More precisely, we are focusing here on a multilayer networks in which the different layers share the same nodes but edges belonging to different types. We built a four-layer multilayernetwork of biological interactions. In all the layers, the nodes correspond to genes or proteins, which are here considered similarly and named by their approved symbols in the HUGO Gene Nomenclature Committee (HGNC) database[20]. The proteinprotein interactions (PPI) network layer was built by merging interactions from *Lit-BM*, *Hi-Union* (downloaded from *HuRI* [21]) and *APID* [22] databases. The pathway network layer was downloaded from Reactome[18]. Reactome provides a dataset of protein relationships inferred from pathway data. The protein relationships were inferred between reactants, polymers and all protein components of complexes (containing less than four components) involved in Reactome pathways. We built the co-expression network layer from RNA-Seq data publicly available on the Human Protein Atlas[23]. We applied a Weighted Gene Correlation Network Analysis (WGCNA) on the Human Protein Atlas RNA-seq data using the WGCNA R package[24]. We selected a soft threshold power of 7 and exported the network with an adjacency threshold of 0.5. The molecular complex network layer was built by merging data from the *CORUM* database[25] and the Human Protein Complex Map[26], using the R package OmniPathR[27]. We uploaded the four network layers composing the multilayernetwork to the NDEx platform[28]. Network layer sizes and densities are detailed in Supplementary Table S3. We used Cytoscape[29] to visualize the networks.

### 2.3 Community identification with iterative Random Walk with Restart (itRWR)

Our community identification strategy is based on the Random Walk with Restart (RWR) algorithm. The RWR algorithm simulates a particle walking randomly in the network, starting from one or more specific nodes called the seed(s). At each step, the particle randomly either walk from a node to one of its neighbors or restart in the seed(s).

The idea behind RWR is that the network nodes that are more likely to be visited during this random walk are more relevant to the seed(s) than those that are less likely to be visited. A iterative version of the RWR has been proposed by Macropol et al. [30]. Following this idea, we here implemented a novel approach, called iterative Random Walk with Restart (itRWR), to identify communities around seed node(s) in a multilayer network. To this goal, we extended our previous RWR approaches able to extract communities from multilayer network[31,32]. The itRWR first apply RWR to an initial set of seed(s). Then, at each iteration, the top-scored node is added to the set of seeds and the RWR is run again with this new set of seeds. The algorithm stops when a given number of iterations is reached. The itRWR implementation is based on the MultiXrank Python package[32] and is available as a Python package on GitHub (https://github.com/anthbapt/itRWR).

We applied itRWR systematically to each of the 67 PA diseases to extract their communities from the multilayer network described in section 2.2. For each disease, we used its associated gene(s) as initial seed(s). The RWR parameters used to run the itRWR algorithm are described in Supplementary Table S4. The itRWR algorithm was run with 100 iterations, leading to communities of size *100 + n*, with *n* being the initial number of seed(s) for a given disease (Supplementary File S2). For the sake of comparison, we also assessed the communities obtained with 50 and 150 iterations of the itRWR algorithm.

We performed biological annotation enrichment analyses on the community obtained with 100 iterations of itRWR for each disease with *g:Profiler* (version `e108_eg55_p17_0254fbf`)[33], using all the genes belonging to the community as input. We performed this enrichment analysis using the Gene Ontology[17] Biological Processes and Cellular Components, and Reactome pathway[18] annotations. The background used for the statistical tests was restricted to the set of annotated genes contained in the multilayer network. P-values were corrected for multiple testing using the Benjamini-Hochberg procedure[34] and the significance threshold was set at 0.05 (Supplementary File S7).

### 2.4 Community clustering

We computed the similarity between every pair of disease communities by computing the Jaccard index on the gene sets belonging to the two communities. The Jaccard Index is the ratio between the number of common elements between two sets (i.e., the number of genes in the intersection of the two communities) and the total number of elements in the two sets (i.e., the number of genes in the union of the two communities). We gathered the Jaccard Index values in a 67 × 67 similarity matrix, which we then converted into a distance matrix (by subtracting Jaccard Index similarities from 1). We next clustered the communities using hierarchical clustering of the distance matrix with the “average” method. We obtained a dendrogram of diseases. We set a cutoff at 0.7 on the dendrogram to retrieve clusters of diseases. We kept only the clusters containing at least 3 disease for further interpretation, and thereby obtained 6 clusters, each grouping three to 20 disease communities (see Supplementary File S3 and S4 for lists of genes and diseases in each cluster).

The clusters were generated from disease communities computed with 100 iterations of the itRWR algorithm. To assess how the choice of iteration number in the itRWR algorithm affects the clustering results, we compared the clusters generated from 100 iterations of itRWR with those from 50 and 150 iterations. We used the Rand Index for this comparison, a measure that evaluates the similarity of two clustering solutions by considering how disease pairs are grouped across different cluster sets. The Rand Index is calculated by dividing the number of disease pairs that are clustered together in two different sets of clusters by the total number of potential disease pairs. A higher Rand Index indicates greater agreement in disease clustering between the two sets, reflecting more consistent clustering outcomes. Additionally, we tested an alternative cutoff of 0.5 on the dendrogram obtained by clustering the disease communities obtained from 100 iterations of itRWR.

### 2.5 Cluster enrichment analyses

We performed statistical enrichments using classical over-representation analyses with hypergeometric tests (via inhouse implementation or via *g:Profiler* [33]). Appropriate backgrounds were selected for each analysis, and p-values were systematically corrected for multiple testing.

#### 2.5.1 Enrichment in HPO phenotypes

We used the PyHPO package[35] to fetch HPO phenotypes and their associated ORPHANET diseases. We then performed an HPO phenotype enrichment analysis on each of the 6 community clusters. We applied a hypergeometric test to assess HPO phenotype enrichments in each cluster based on the HPO phenotypes associated with all the diseases belonging to the cluster. We used the total number of ORPHANET diseases associated with at least one HPO phenotype as background (i.e, 4 262 diseases associated with 1 364 different phenotypes). P-values were corrected using the Benjamini-Hochberg procedure[34] and the significance threshold was set at 0.05 (Supplementary File S9).

#### 2.5.2 Enrichment in biological annotations

We performed biological annotation enrichment analyses on each of the six community clusters with *g:Profiler* (version `e108_eg55_p17_0254fbf`)[33], using the union of the genes belonging to the disease communities of each cluster as input. We performed these enrichment analyses using the Gene Ontology[17] Biological Processes and Cellular Components, and Reactome pathway[18] annotations. The background used for the statistical tests was restricted to the set of annotated genes contained in the multilayer network. P-values were corrected for multiple testing using the Benjamini-Hochberg procedure[34] and the significance threshold was set at 0.05 (Supplementary File S8). We filtered the enrichment results using the *orsum* Python package[36]. *orsum* filters enrichment results by selecting more general and significant annotation terms as representatives for their less significant child terms. In addition, *orsum* allows integrating and representing enrichment results obtained in different analyses, in our case enrichments results obtained for the six community clusters. In *orsum*, we used the default parameters except the maximum representative term size (maxRepSize) that we set to 2 000 in Reactome pathway analyses.

#### 2.5.3 Enrichment in physiological aging annotations

We performed enrichment analyses on each of the six community clusters to reveal enrichments in genes associated with physiological aging. To this goal, we used a list of 307 aging-related genes from the GenAge database[37]. From this list, we kept only the genes with a direct association with aging, i.e., we removed the genes presenting the association types “upstream”, “downstream”, “functional” and “putative”. The final list of GenAge aging genes contains 107 genes. In order to analyse the enrichments of our six community clusters in GenAge aging genes, we performed a hypergeometric test. P-values were corrected using the Benjamini-Hochberg procedure[34] and the significance threshold was set at 0,05 Supplementary File S5).

We performed similar enrichment analyses using lists of genes up and down-regulated during aging in five different human tissues extracted from the study of Irizar et al. [38]. The number of genes up-regulated and down-regulated in each tissue is detailed in the Supplementary File S6. The lists of genes differentially expressed during aging in blood and skin were produced by the study of Irizar et al. The lists of genes differentially expressed during aging in brain, muscle and breast were extracted by Irizar et al. from three other publications (respectively[39],[40] and[41]). We mapped the Ensembl identifiers to The HUGO Gene Nomenclature Committee (HGNC) database[20]. We performed hypergeometric tests for genes up-regulated and down regulated in each of our six community clusters. P-values were corrected using the Benjamini-Hochberg procedure[34] and the significance threshold was set at 0.05 (Supplementary File S5).

### 2.6 Implementation

The itRWR algorithm is available as a Python package at https://github.com/anthbapt/itRWR. The pipeline developed for this study, including the identification of communities with itRWR, the clustering of communities, and the cluster enrichment analyses, is available on GitHub at https://github.com/CecileBeust/PA_Communities.

## 3 Results

### 3.1 Selecting premature aging diseases and their associated genes

The workflow of our study is available in Figure 1. To systematically identify Premature Aging (PA) diseases, we selected the HPO annotation “*HP:0007495: prematurely aged appearance*” and its 12 descendant phenotype annotations in the ontology tree (Figure 1a, Supplementary Table S1). We then selected, from ORPHANET, the diseases associated with at least one of these HPO phenotypes and associated with at least one gene. Overall, this led to the identification of 67 PA diseases associated with a total of 132 genes, ranging from one to 20 associated genes per PA disease (Supplementary Table S2). We observed that the majority of the genes are mutated in only one disease (Figure 1b).

**Figure 1:**
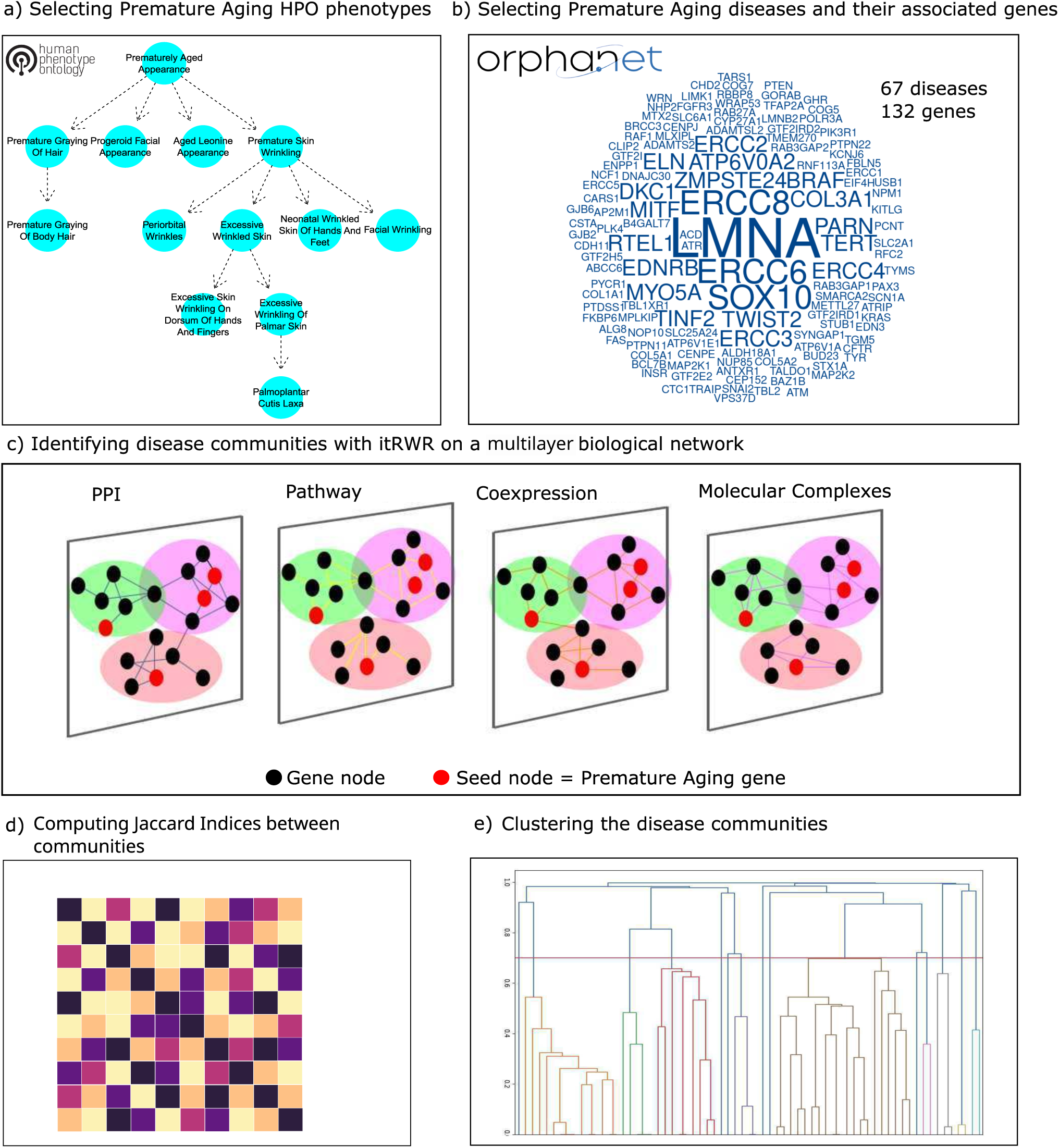
Workflow of the study. Overview of the workflow developed to systematically identify the cellular functions associated with premature aging (PA) diseases a) 13 PA phenotypes are selected among HPO phenotypes [15]. b) Based on the 13 PA phenotypes, 67 PA diseases and their 132 associated genes were selected from ORPHANET. The size of the gene name in the word cloud corresponds to the number of diseases in which the gene is mutated. c) Disease community identification with iterative Random Walk with Restart (itRWR). For each disease independently, the disease-associated gene(s) were used as seed(s) in the itRWR algorithm. itRWR is applied to a multilayer network composed of four layers: protein-protein interactions, pathways, molecular complexes and co-expression. In the figure, for the sake of visualization, only three toy disease communities are represented (in pink, salmon, and green). d) Jaccard Indices werecomputed between the gene sets of each pair of disease communities. e) A clustering algorithm was applied on the Jaccard Index matrix, and clusters of disease communities were identified with a cutoff on the dendrogram.

The selected diseases include prototypical PA diseases such as Hutchinson-Gilford Progeria Syndrome (HGPS, ORPHANET code: 740), characterized by osteoporosis, lipodystrophy and atherosclerosis[4]. HGPS is caused by specific mutations in the *LMNA* or *ZMPSTE24* genes. These mutations lead to the production of misprocessed Prelamin A isoforms and a loss of the integrity of the nuclear lamina. This loss of integrity impairs several major cellular functions, such as DNA repair, cell cycle, transcription, splicing, translation, or apoptosis[1]. Another example of selected PA disease is Ataxia-telangiectasia (ORPHANET code: 100), which is characterized by cerebellar degeneration, telangiectasia (dilatation of small surface blood vessels), immunodeficiency, cancer susceptibility and radiation sensitivity[42]. It is caused by mutations in the *ATM* gene, a DNA damage sensor. Ataxia Telangiectasia belongs to the DNA repair deficient syndromes, which were among the firsts PA diseases to be deciphered. A last example is the Ehlers-Danlos syndrome (ORPHANET code: 287). It is characterized by joint hyper-mobility, skin hyper-elasticity and generalized fragility of soft tissue[43]. It can be caused by mutations in several collagen genes (*COL1A1, COL5A1, COL5A2*) impacting the extracellular matrix organization. Hence, these three diseases described here as examples all share a PA phenotype, but are associated with the perturbation of initially distinct molecular mechanisms.

In order to study the cellular functions perturbed in PA diseases, we first performed a classical functional enrichment analysis of the 132 genes associated with the 67 PA diseases (Material and Methods). Most of the significant annotations are related with nuclear processes (Supplementary File S1). We hypothesized that such classical enrichment analysis, focusing solely on the genes directly linked to PA diseases, does not allow revealing a comprehensive landscape of deregulated cellular functions. We hence decided to expand our analysis to study the biological network neighbourhood of the disease-associated genes.

### 3.2 Extracting PA disease communities from a biological multilayer network with iterative Random Walk with Restart

To elucidate the cellular functions underlying PA diseases, we systematically extracted the PA disease communities from a multilayer biological network. To this goal, we developed and applied an iterative Random Walk with Restart (itRWR) algorithm (Materials and Methods). The itRWR algorithm explores a multilayer network starting from nodes of interest, called the seeds. For each PA disease, we designated its associated gene(s) as the seed node(s), and executed the itRWR algorithm (Figure 1c). Following this approach, we obtained 67 disease communities (Supplementary File S2).

Each community, gathering the multilayer network neighbourhood of the disease-associated gene(s), provides information into the cellular functions that might be disrupted in the disease. For instance, the community obtained for Hutchinson-Gilford Progeria Syndrome (HGPS) contains 102 gene nodes (two seed nodes and 100 nodes retrieved by itRWR, Supplementary File S2). Its enrichment analyses depict a significant over-representation of genes involved in DNA repair and other nuclear processes (Material and Methods, Supplementary File S7). The community identified for Ataxia-telangiectasia, using its mutated gene *ATM* as seed in the itRWR, contains 101 gene nodes. This disease community displays similar functional enrichments as the HGPS community (Supplementary File S7). Finally, the community obtained for the Classical Ehlers-Danlos syndrome contains 103 gene nodes. In this case, the results reveal a significant over-representation of genes involved in extracellular signaling pathways as well as cell adhesion and cell migration functions (Supplementary File S7).

To further explore whether Ataxia-telangiectasia, HGPS, and Classical Ehlers-Danlos syndrome share perturbations of similar cellular functions, we compared the gene nodes belonging to their respective disease communities (Figure 2). We observed a high degree of overlap between the HGPS and Ataxia-telangiectasia communities, with 87 common genes over 102 and 101 community gene nodes for HGPS and Ataxia-telangiectasia, respectively. These common gene nodes predominantly encode proteins related to DNA repair and response to DNA damage, such as proteins of the DNA non-homologous end joining complex (e.g., *XRCC4-6*).

**Figure 2:**
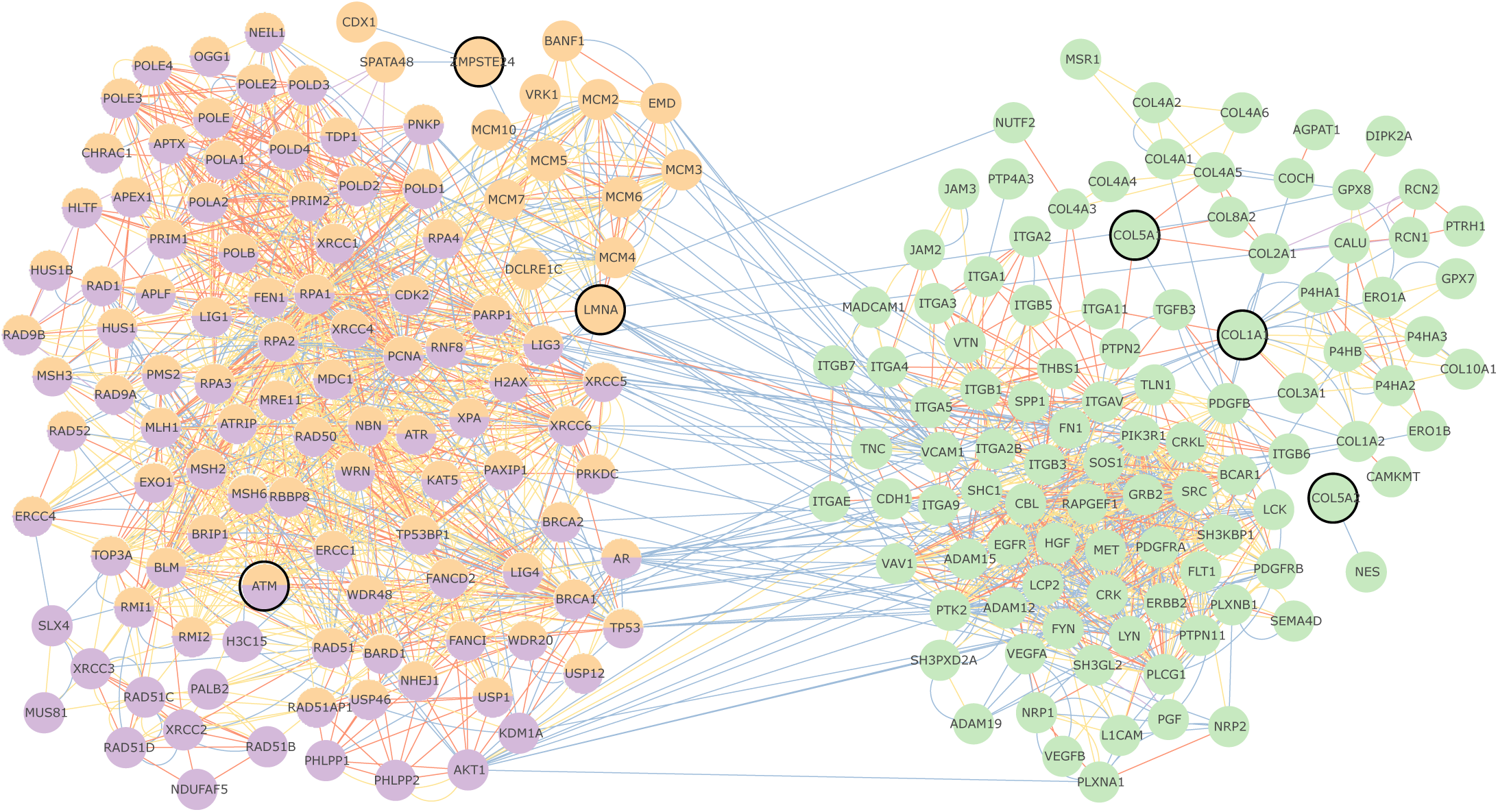
Disease communities of Hutchinson-Gilford Progeria Syndrome (in orange, 102 nodes, seeds: *LMNA* and *ZMPSTE24*), Ataxia-telangiectasia (in violet, 101 nodes, seed: *ATM*), and Classical Ehlers-Danlos syndrome (in green, 103 nodes, seeds: *COL1A1*, *COL5A2*, *COL5A1*). The communities were identified using itRWR on a multilayer network composed of four layers of biological interactions. The seed node(s) used for each disease are circled in black. Edge colors correspond to the interaction layer of the multilayer network: Protein-protein interactions (blue), pathways (yellow), molecular complexes (orange), co-expression (purple).

A small number of nodes are retrieved only in one of the two disease communities. For example, the phosphatases *PHLPP1, PHLPP2* are found only in the Ataxia-telangiectasia community. Contrarily, different proteins of the *MCM* family, which are involved in DNA replication, as well as proteins involved in the nuclear envelope architecture, such as *EMD*, are found only on the HGPS community (Figure 2).

Interestingly, the gene nodes of these two disease communities show no overlap with those of the Ehlers-Danlos syndrome community (Figure 2), indicating a markedly distinct node composition. The Ehlers-Danlos syndrome community mainly contains genes encoding proteins involved in tyrosine-kinase signaling pathways, e.g. *SRC* or *ERBB2*. Some of them are growth factors, like *EGFR*, which is known to be involved in cell proliferation. The community also contains several collagen proteins. Collagens are known to act as ligands of several receptors including the tyrosine-kinase receptors *DDR1* and *DDR2* [44].

Overall, the three diseases used here as example seem to be associated with the perturbation of two different cellular functions, demonstrated by the grouping of the disease communities into two distinct subnetworks. In the next section, we extended the comparison to systematically assess the overlap between the disease communities extracted for the 67 PA diseases.

### 3.3 Clustering disease communities defines the molecular landscape of PA diseases

We hypothesized that PA disease communities with a large number of overlapping gene nodes might reveal shared disease-associated molecular mechanisms. We hence systematically compared the overlaps between the communities associated with the 67 PA diseases. To this goal, we computed the Jaccard Index between the gene sets composing each pair of disease communities (Figure 1d,e). The resulting matrix was then used to build a clustermap (Figure 3, Materials and Methods). After setting a cutoff of 0.7 in the dendrogram and filtering out the clusters composed of less than three disease communities, we obtained six clusters containing between three to 20 disease communities (Figure 3, Supplementary Files S3 and S4). Importantly, the obtained clusters are robust to changes in the number of iterations used in the itRWR algorithm to identify the disease communities. Indeed, the set of clusters obtained using 100 iterations in itRWR clustering is similar to the sets of clusters obtained using 50 iteration and 150 iteration (Rand indices equal to 0.74 and 0.96, respectively, Material and Methods, Supplementary Figure S1 and S2).

**Figure 3:**
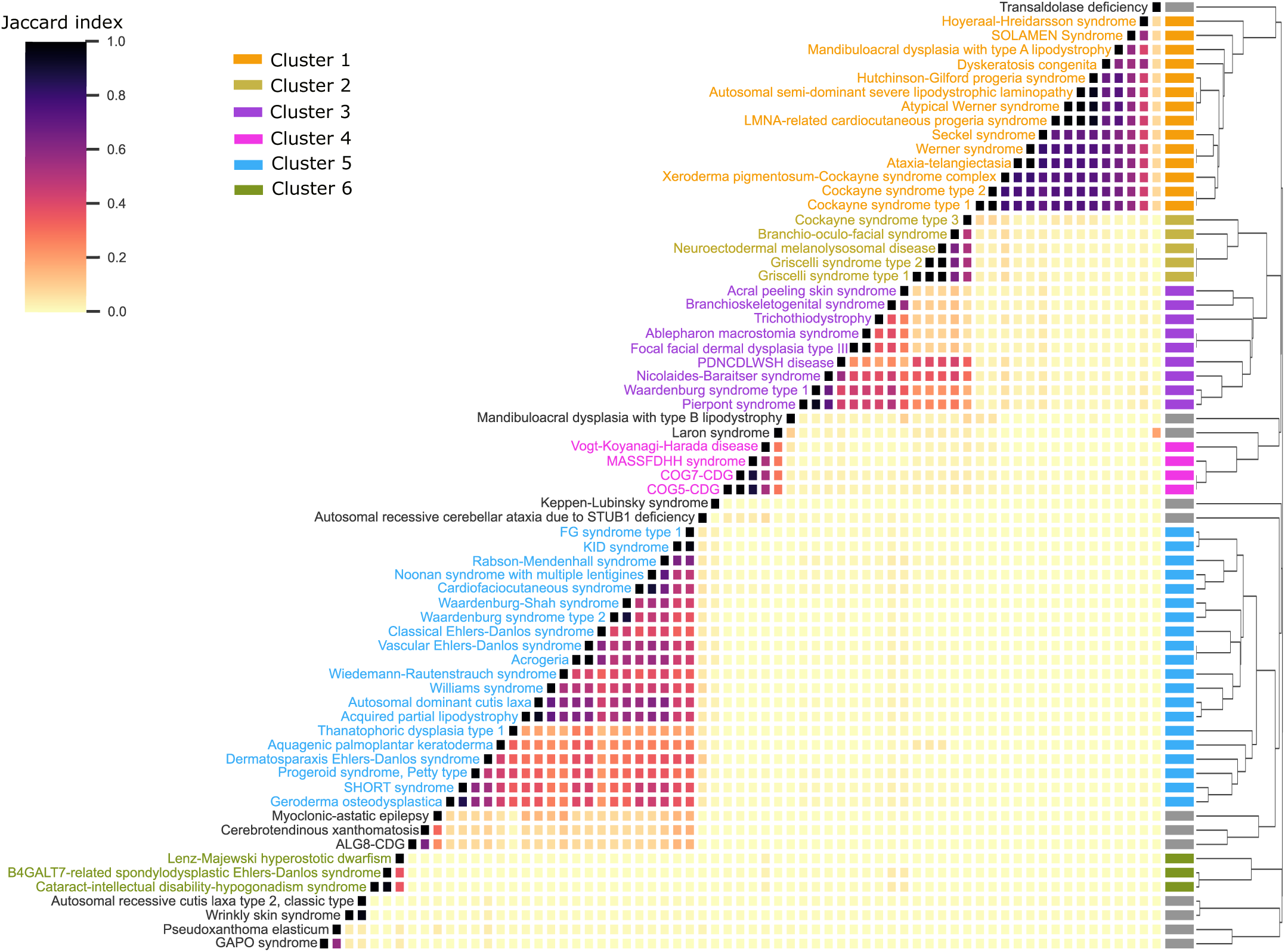
Clustering of the premature aging disease communities. Pairwise Jaccard indices were computed between the gene sets associated with each pair of disease communities. Then, the resulting matrix was used as input for hierarchical clustering. In the clustermap, the color gradient represents the similarities between communities (darker cells correspond to closer communities according to their Jaccard Indices). The clustering uses the average method and outputs a dendrogram. Clusters were determined by setting a cutoff value at 0.7 on the dendrogram. We kept the six clusters that contained at least three disease for downstream analyses. MASSFDHH: Moyamoya angiopathy-short stature-facial dysmorphism-hypergonadotropic hypogonadism syndrome. PDNCDLWSH: Peripheral demyelinating neuropathy-central dysmyelinating leukodystrophy-Waardenburg syndrome-Hirschsprung disease. SOLAMEN syndrome: Segmental outgrowth-lipomatosis-arteriovenous malformation-epidermal nevus syndrome

We next assessed the enrichments in cellular localizations, biological processes, and pathways, in each cluster, using the annotations of the genes within these clusters (Materials and Methods, (Supplementary File S8)). At a coarse-grained level, we observed that the clusters 1, 2, & 3 have similar annotation profiles for cellular localization (Figure 4a). Indeed, the genes composing these three clusters are significantly enriched in nucleus, nucleoplasm and chromosome localizations. This nuclear localization is also reflected at the level of enriched pathways and processes, which are overall related to mitosis, cell cycle, gene expression and metabolic processes (Figure 4b, c). In contrast, the clusters 4, 5, & 6 are enriched in non-nuclear localization, including membrane, cytoplasm, and/or cell periphery localization (Figure 4a). This is also reflected by the enrichments in signal transduction and cell communication identified in these clusters (Figure 4b, c). Furthermore, we examined the cluster enrichments in phenotypes, using the HPO annotations of the diseases belonging to each cluster (Materials and Methods, Supplementary File S9). It should be noted that the HPO phenotype “prematurely aged appearance” is significant in all clusters. This is in line with the initial criteria used to select PA diseases. Indeed, PA diseases were selected as associated with this specific HPO term or any of its descendant terms. Our analysis did not identify any other distinct patterns in phenotype enrichments (Figure 4d).

**Figure 4:**
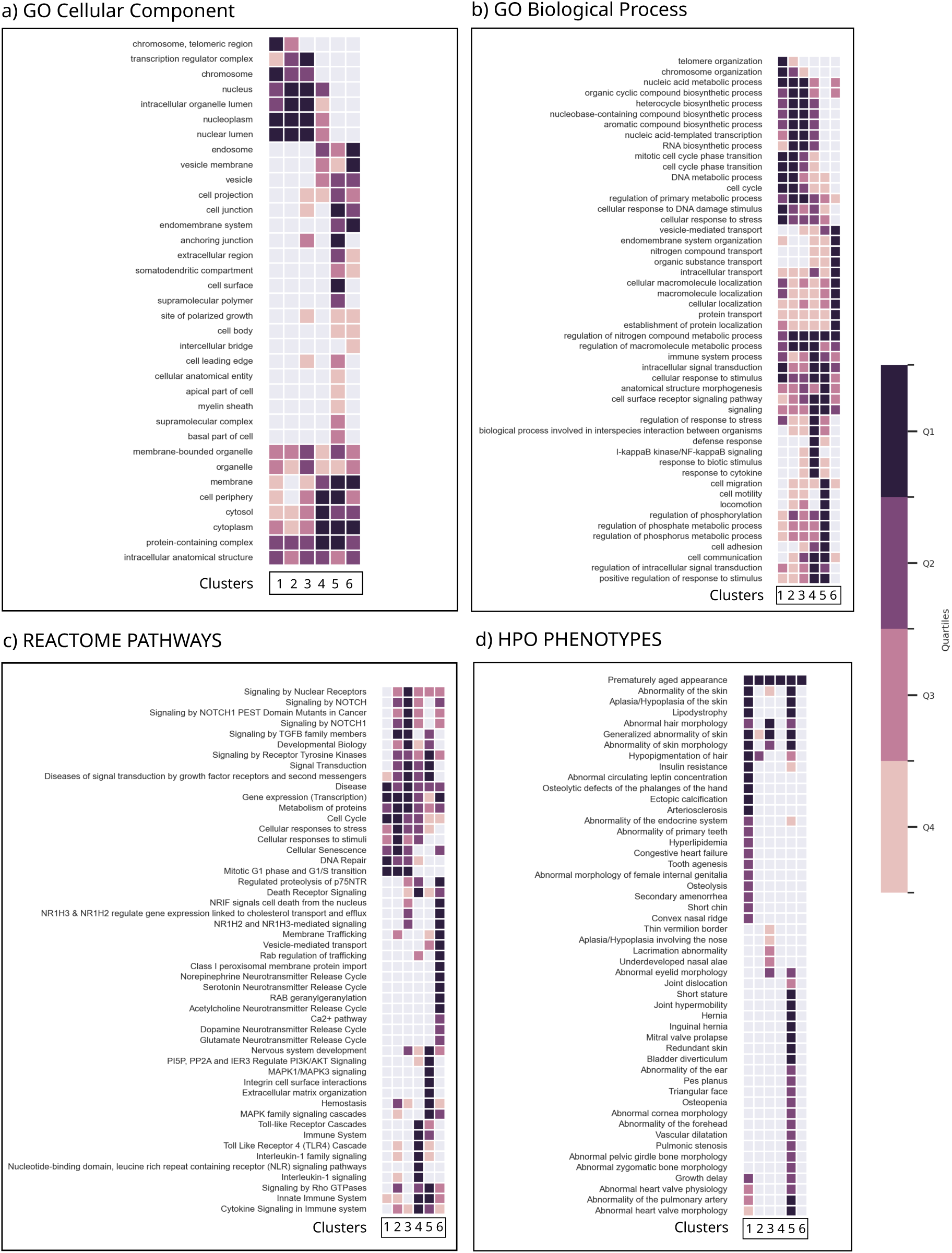
Cluster enrichments in (a) Cellular Components, (b) GO Biological Processes, (c) Reactome pathways, and (d) HPO phenotypes. For GO Biological Process, Cellular Components and Reactome pathways, enrichments were computed with g: Profiler using the cluster gene lists as inputs and filtered using orsum. For HPO, a hypergeometric test was applied using the cluster disease lists as input. In both cases, significant annotations were selected with a threshold set at 0.05 on the corrected p-values. The color gradient reflects the quartile of the annotation significance.

We next focused more deeply on the annotation enrichments of each cluster (Supplementary Files S8 and S9). Cluster 1 gathers 14 diseases and a total of 232 genes. As stated previously, the genes belonging to this cluster have significant enrichments in nuclear and chromosome localization annotations, similar to Clusters 2 & 3. However, Cluster 1 displays an additional very strong enrichment in telomeric regions of the chromosomes. At the biological process and pathways levels, this is reflected by significant enrichments in telomere and chromosome organization processes. The most significant cellular functions of Cluster 1 are DNA metabolic process and DNA repair. The HPO phenotype enrichment analysis of the diseases belonging to Cluster 1 reveals skin and hair abnormalities, bone and cardiovascular defects, lipodystrophy, and growth delay. The second cluster contains five diseases and a total of 156 genes. At the functional level, its most over-represented annotations are related to mitotic G1 phase and G1/S transition. A significant HPO enrichments is *Hypopigmentation of hair*. Cluster 3 contains nine diseases and a total of 282 genes. It displays similar nuclear localization enrichments as Clusters 1 & 2 but with an additional significant enrichment in genes annotated for the transcriptor regulator complex. The most significant functional enrichments are related to transcription regulation. Significantly enriched Reactome pathways point to signal transduction, including signaling by nuclear receptor, Notch and TGFB. HPO enrichments exhibit phenotypes also retrieved in other Clusters, such as hair and skin abnormalities, and some specific phenotypes, for instance ocular lacrimation abnormalities. In summary, despite their shared enrichments in nuclear localization, biological processes and pathways, the genes belonging to Clusters 1, 2, & 3 exhibit subtle differences in their cellular function enrichments.

Clusters 4, 5, 6 display enrichment patterns different from the first three clusters (Supplementary File S8 and S9). Cluster 4 is composed of four diseases and 138 genes. The genes belonging to Cluster 4 are significantly annotated with cytoplasmic localizations. At the level of GO Biological Processes, the enriched annotations are mainly related to signal transduction, inflammation, and immune system processes, with the strongest enrichments being in the *I*-*κ*B Kinase/NF-*κ*B signaling. The Reactome pathways enriched in this cluster are Toll-like Receptor cascades, death signaling and immune system related pathways. Cluster 5 is the largest cluster. It contains 20 diseases and 466 genes. The cluster genes are enriched in membrane and cell periphery localizations, but also displays significant enrichments in anchoring junction and extracellular region. These subcellular localizations are reflected at the level of biological process and pathway that reveal enrichments in cell adhesion and migration, extracellular matrix organization and integrin cell surface interactions, and signaling with receptor tyrosine kinases. At the phenotype level, many significant HPO terms are the same as the terms significant in Cluster 1, such as hair and skin abnormalities, and lipodystrophy. But additional phenotypes, such as joint mobility defects, are retrieved only in the Cluster 5. Finally, the last cluster, Cluster 6, contains only three diseases and 150 genes. These genes are enriched in localization annotations related to endosome and vesicle membrane, reflected in processes and pathways enrichments in vesicle-mediated transport and membrane trafficking.

In summary, we observed that the Clusters 1, 2, & 3 display similarly enriched nuclear localizations and processes and pathways related to nuclear functioning. But they also display subtle differences in their cellular functions. Indeed, Cluster 1 is more significantly associated with DNA repair mechanisms, Cluster 2 with cell cycle, in particular mitosis G1/S phases, and Cluster 3 with transcription regulation. Of note, a large majority of the PA diseases considered as classical progeroid syndrome[45], such as Hutchinson-Gilford Progeria or Werner Syndromes, belong to Cluster 1. The three last clusters (Clusters 4, 5, & 6) are associated with different and non-nuclear molecular mechanisms. Cluster 4 gathers inflammation and immune system pathways; Cluster 5 is associated with cellular communication; Cluster 6 is related with vesicle-mediated transport. Overall, the six clusters hence reveal a landscape of the molecular mechanisms associated with PA diseases.

It is to note that similar clusters and cluster functional enrichments are retrieved with an alternative cutoff of the dendrogram at 0.5 (Supplementary Figure S3 and File S8bis). In addition, we repeated our workflow with a classical (non-iterative) RWR, a global partitioning of the multilayer network with MolTi 2.0 [14], and computing the shortest paths distances between disease genes. All these alternative network exploration strategies were able to highlight some disease relationships, collectively revealing that the signal is present in the multilayer network. However, the itRWR amplified the detection of this signal by revealing larger clusters of diseases (Supplementary Text S1).

### 3.4 Linking the molecular landscape of PA diseases to Physiological Aging

In order to identify potential links between premature and physiological aging, we assessed the cluster enrichments in genes related to physiological aging. We first tested whether the clusters are enriched in physiological aging genes extracted from the GenAge database[46] (Material and Methods). This analysis revealed that all clusters except cluster 6 are significantly enriched in GenAge genes (corrected p-values from 10^−14^ to 10^−03^, Figure 5, Supplementary File S5). Next, we tested the cluster enrichments in genes up and down-regulated during physiological aging in five different human tissues (blood, skin, brain, muscle and breast, Materials and Methods). We observed that Cluster 5 is enriched in genes up-regulated in skin during physiological aging (corrected p-value = 4.9 × 10^−04^). This is consistent with the set of PA diseases gathered in Cluster 5, such as different types of Ehlers-Danlos syndromes or the Autosomal dominant cutis laxa, which are mainly characterized by skin abnormalities. In Clusters 4 & 5, we noticed a strong enrichment in genes up-regulated in the brain during physiological aging. Finally, Clusters 2 & 3 display significant enrichments in genes up-regulated in muscle and down-regulated in breast during physiological aging. Cluster 6 does not present any significant enrichment in genes associated with physiological aging, even if some of the cluster genes are deregulated, in particular down-regulated in brain tissue during physiological aging (Supplementary File S5).

**Figure 5:**
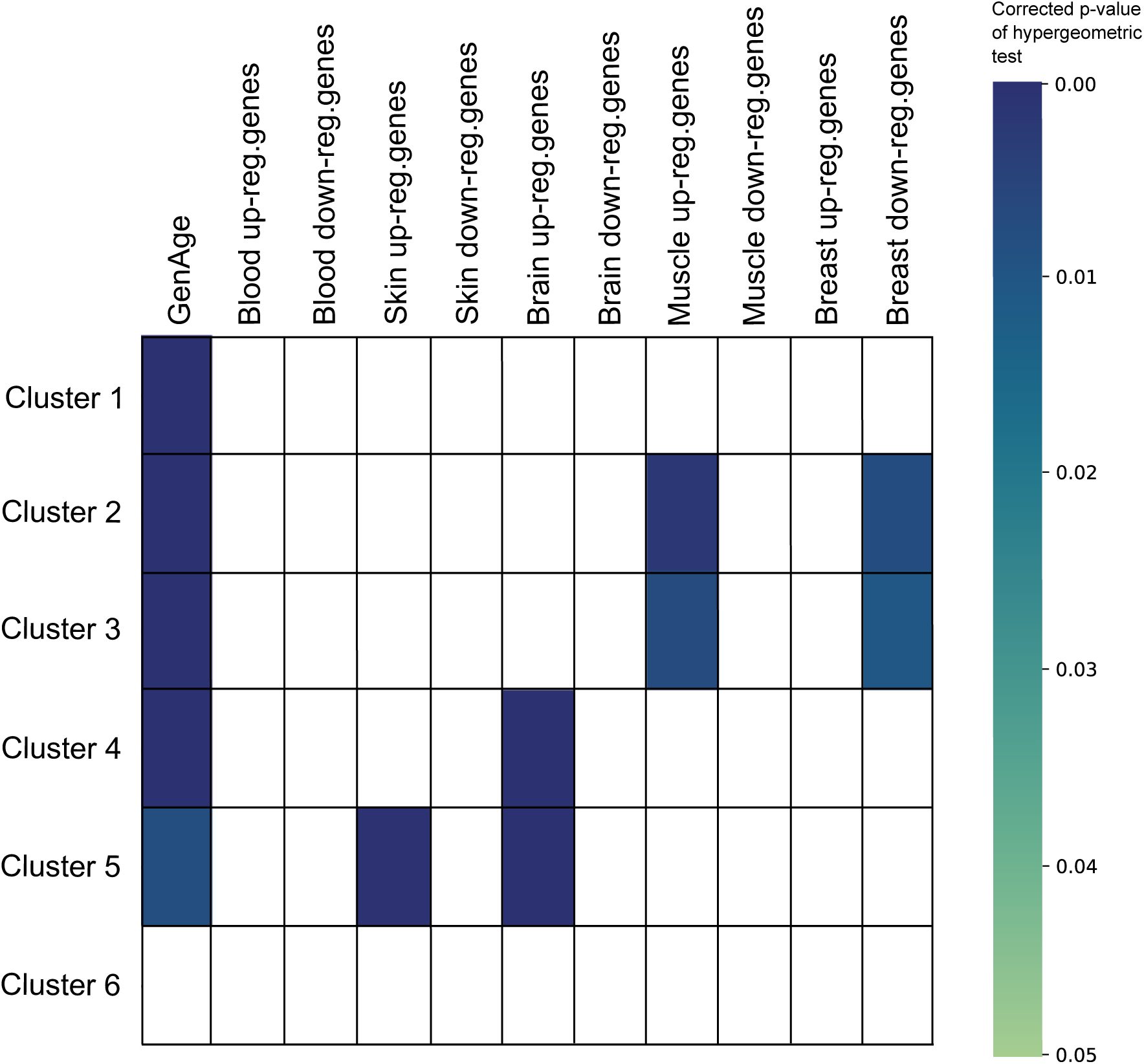
Cluster enrichments in physiological aging genes. The first column represents the enrichment results in physiological aging genes extracted from the GenAge database [46]. The other columns represent enrichment results of clusters in genes up and down-regulated during physiological aging in several tissues from the study of [38]. The colors represent the p-value of the hypergeometric test.

## Discussion

In this study, we provide a detailed overview of the molecular mechanisms involved in PA diseases through a systematic and data-driven functional analysis of genes associated with these conditions. This landscape expands the scope of the molecular mechanisms previously reviewed for PA diseases. Indeed, to the best of our knowledge, these molecular mechanisms were so far only studied from a literature review perspective. Recent literature reviews broadly classify PA diseases into three groups corresponding to defects in nuclear architecture, DNA repair, and other processes[1,5,6]. The data-driven molecular landscape we propose here groups the PA diseases into six clusters primarily annotated for DNA repair, cell cycle, transcription regulation, inflammation, cell communication, and vesicle-mediated transport.

Overall, the first three clusters exhibit a close functional relationship, as they are all associated with nuclear processes. Specifically, Cluster 1 predominantly pertains to DNA repair mechanisms, Cluster 2 is more significantly linked to cell cycle regulation, and Cluster 3 primarily involves transcription regulation. These mechanisms represent interconnected nuclear processes, which tend to become dysregulated in a cascade-like fashion when any one of them is impaired.

Regarding PA diseases associated with defects in the nuclear envelope architecture, typically caused by mutations in lamin A or proteins interacting with lamin A, our findings reveal their belonging to Cluster 1 rather than the formation of an isolated cluster. Cluster 1 most significant annotations pertain to DNA repair processes. The maintenance of nuclear envelope integrity is closely tied to DNA repair, as evidenced by the intra-nuclear accumulation of progerin (the truncated isoform of prelamin A in Hutchinson-Gilford Progeria Syndrome) leading to impaired repair of double-strand DNA breaks and delayed recruitment of ATM proteins at breakage sites[47]. While existing evidence underscores such functional link between nuclear envelope architecture and DNA repair, we hypothesize that the discovery of novel interactions and the expansion of biological networks may lead to the division of Cluster 1 into two subclusters, potentially reflecting functional nuances among these PA diseases.

The remaining three clusters predominantly relate to non-nuclear cellular functions. Cluster 5 encompasses genes and proteins involved in cell communication and extracellular processes, including those implicated in Ehlers-Danlos syndromes, which affect connective tissues. Cluster 4, characterized by its association with inflammation, includes genes related to glycosylation disorders. Notably, Cluster 6, primarily annotated for vesicle-mediated transport, represents a cellular function that, to our knowledge, has not been reviewed in the context of PA diseases. This underscores a potentially under-explored area in PA research, suggesting new avenues for investigation.

Overall, our data-driven approach has allowed us to systematically classify PA diseases and broaden our understanding of their molecular perturbations. As discussed previously, the expansion of biological interaction networks is expected to further refine our clusters. Particularly, larger clusters such as Clusters 1 and 5, which currently encompass a broad range of PA diseases, may eventually be subdivided into more specific subclusters. Furthermore, we expect that certain diseases that are currently not part of any cluster due to the prerequisite of a minimum of three closely related diseases for cluster formation, could form new clusters as additional interaction data becomes available. We can also note that our methodology focused on clustering disease communities, thereby spotlighting the shared cellular functions among PA diseases. An alternative approach could be to examine disease-specific subnetworks. For instance, in Figure 2, we can pinpoint nodes unique to the HGPS disease community. The HGPS-specific nodes include *EMD*, which codes for lamin A-interacting protein Emerin, and various DNA replication factors. This kind of targeted analysis could yield deeper insights into the unique molecular pathways and interactions characteristic of each PA disease. It is also important to note that the data used as input in our analyses are genetic and biological network data. The availability of additional omics datasets for PA diseases, in particular transcriptomics datasets, would be of high interest to validate and extend our finding.

We systematically selected all the diseases associated with a PA phenotype using HPO[48] and ORPHANET[16] databases. This approach gathered a wide spectrum of diseases with various clinical presentation and molecular pathogenesis. Our set of PA diseases include classical progeroid syndromes, such as Hutchinson-Gilford Progeria Syndrome, which are unambiguously considered as PA syndromes in the literature, but also other diseases which belonging to the category of progeroid syndrome or PA diseases might be debatable. For instance, the premature aging phenotype present in Griscelli syndrome patients is “premature graying of hair”, and the classification of this syndrome as PA might be questioned[49]. In addition, we also identified some questionable gene-disease associations. For instance, in our study, Acrogeria is grouped in Cluster 5 alongside Ehlers-Danlos PA diseases. This clustering stems from Acrogeria’s association with mutations in *COL3A1* in ORPHANET. However, this indication does not consider various pieces of evidence linking Acrogeria to disease-causing variants in *LMNA* and *ZMPSTE24* [50,51]. Factoring in these associations might shift Acrogeria into Cluster 1. Another example are Cockayne Syndromes that are split in clusters 1 and 2. Two main Cockayne Syndrome types exist, CSA and CSB, caused by disease-causing variants in *ERCC6* and *ERCC8*. However, databases such as ORPHANET and OMIM[52] have documented extremely rare phenotypes as Cockayne Syndromes related to other variants in *ERCC1* and *ERCC4*. Misdiagnoses, whether clinical or molecular, are a possibility that can introduce errors into database entries. The algorithm we developed in this study incorporates these data, although they may not be entirely relevant, potentially affecting the accuracy of our findings. Overall, an alternative to our systematic data-driven strategy could have been to select a set of diseases based on literature reviews or based on expert knowledge. However, in order to ensure unbiased and reproducible studies, we preferred not to filter a priori the diseases.

Even if the global incidence of premature aging diseases is low (estimated 1:4 000 000 for Hutchinson-Gilford Progeria Syndrome, 1:50 000 for progeroid syndromes[3], for instance), rare diseases represent a significant burden for patients and their families[53]. Our data-driven classification could lead to a deeper understanding of PA diseases, with compelling implications for diagnosis and treatment. Firstly, unraveling the molecular underpinnings of these syndromes can improve diagnosis and pave the way for targeted therapies. More specifically, multilayer interaction data could aid in identifying previously unknown biological connections and crucial disease biomarkers, essential for clinical trials but often missing in PA and rare diseases. Secondly, categorizing these diseases into groups with shared cellular dysfunction can open up new avenues for treatment strategies. For instance, therapeutic molecules targeting different diseases belonging to the same cluster could be developed once the common pathophysiological mechanism has been validated. Moreover, innovative therapies, akin to those for rare cancers, might be adapted for non-oncological rare diseases. Preclinical *in vitro* and *in vivo* studies confirming the potential efficacy/safety ratio of a drug and its relevance in multiple indications, might lead to include patients being affected with different phenotypes in early clinical phases. Such a “basket trial” could expedite clinical timelines and be cost-effective[54]. Lastly, a better understanding of PA disease etiology and pathogenesis can shed light on the molecular mechanisms involved physiological aging[49,1].

Aging is a multifaceted process governed by numerous molecular mechanisms, many of which are yet to be fully understood. PA diseases are an important source of information for studying the molecular mechanisms that could also be perturbed during physiological aging[1]. In recent years, knowledge of the links between PA diseases and physiological aging has been increasing. Our approach, which leverages different sources of information to delineate the molecular landscape of PA and connect it to the known and deregulated physiological aging genes, is interesting for hypothesis generation. For instance, our observation that Cluster 5 is enriched in genes up-regulated in physiological skin aging is relevant, as the main phenotypic pattern of PA diseases included in this cluster is a skin defect. Likewise, the association of Cluster 4 with genes that are up-regulated in brain tissue during aging may underscore the role of inflammation as a primary and secondary mechanism in neurological aging.

Our PA molecular landscape was defined based on the exploration of a multilayer interaction network. The study of biological networks in pathological contexts is at the heart of network biomedicine[55]. Network medicine assumption is that molecular components associated with similar diseases are belonging to a close network vicinity[56]. Biological interactions have been demonstrated as important features to predict aging-related genes[57,58,59,60], and network medicine approaches have been successfully used to study aging-related genes and molecular mechanisms[61,62,63,64].

In order to leverage as much as possible the interaction information available in the literature, we consider jointly multiple interaction sources gathered in a multilayer network. Considering multilayer networks is a pertinent way to reduce the incompleteness of individual interaction sources[65]. Multilayer networks represent a formidable source of information to study cellular functioning and identify disease-associated perturbations, but they are large and complex and appropriate exploration tools are mandatory. Community identification is widely use to identify subnetworks of genes or proteins involved in common cellular processes[11]. Different algorithms exist for community identification in multilayer networks[12]. For instance, the ACLcut algorithm is a local community identification approach based on RWR [66]. We proposed here a novel approach called iterative Random Walk with Restart. This novel approach is based on our previous work extending the Random Walk with Restart algorithm to explore multilayer networks[31,32]. We decided to set a maximal number of iterations to obtain disease communities of similar sizes. However, future version of the algorithms could be implemented with a different stopping criteria, for instance by optimizing measures such as the conductance measure in the ACLcut algorithm[66]. Importantly, our itRWR and protocol can be reused to evaluate and compare the molecular bases of any set of diseases.

## Conflict of Interest

AV is employee and shareholder of F. Hoffmann-La Roche Ltd. NL is shareholder of ProGeLife and its subsidiary Calysens. NL is employee (Chief Scientist Rare Disease) at the insternational research institute Servier.

## Author Contributions

Beust C: Conceptualization, Formal Analysis, Software, Original Draft Preparation, Writing, Review & Editing Valdeolivas A: Conceptualization, Original Draft Preparation, Writing, Review & Editing Baptista A: Software, Writing, Review & Editing Brière G: Supervision, Writing, Review & Editing Lévy N: Writing, Review & Editing Ozisik O: Supervision, Software, Writing, Review & Editing Baudot A: Project Administration, Conceptualization, Supervision, Original Draft Preparation, Writing, Review & Editing

## Data Availability

All data and codes produced are available online at:
– Supplementary Materials: https://zenodo.org/records/12516969
– Networks: https://www.ndexbio.org/index.html%23/search?searchType=All&searchString=cecile.beust&searchTermExpansion=false
– Community identification in multilayer networks: itRWR Python Package https://github.com/anthbapt/
itRWR
– Complete analysis workflow: Community identification, clustering, and enrichment analyses https://github.com/CecileBeust/PA_Communities.git

https://www.ndexbio.org/index.html%23/search?searchType=All&searchString=cecile.beust&searchTermExpansion=false

https://github.com/anthbapt/itRWR

https://github.com/CecileBeust/PA_Communities.git

## Acknowledgments

This work received support from the French National Research Agency (ANR-21-CE45-000101), the Association Française contre les Myopathies (AFM), the Excellence Initiative of Aix-Marseille University – A*Midex, a French “Investissements d’Avenir” programme – Institute MarMaRa AMX-19-IET-007, and the European Union’s Horizon 2020 research and innovation programme under the EJP RD COFUND-EJP No 825575. GPT-4 was used to improve syntax and readability.

## Data and code availability

- Data:
  – Supplementary Materials: https://zenodo.org/records/12516969
  – Networks: https://www.ndexbio.org/index.html%23/search?searchType=All&searchString=cecile.beust&searchTermExpansion=false
- Code:
  – Community identification in multilayer networks: itRWR Python Package https://github.com/anthbapt/ `itRWR`
  – Complete analysis workflow: Community identification, clustering, and enrichment analyses https:// github.com/CecileBeust/PA_Communities.git

## Supplementary Materials

### Supplementary Figures

Supplementary Figure S1: Clustering of the premature aging disease communities obtained with 50 iterations of itRWR

Supplementary Figure S2: Clustering of the premature aging disease communities obtained with 150 iterations of itRWR

Supplementary Figure S3: Clustering of the premature aging disease communities obtained with 100 iteration of itRWR, with a cutoff value at 0.5 on the dendrogram

### Supplementary Tables

Supplementary Table S1: Premature Aging HPO phenotypes

Supplementary Table S2: the 67 PA diseases and their 132 associated genes from ORPHANET

Supplementary Table S3: Number of nodes, edges, and densities of the 4 network layers composing the multilayer biological network

Supplementary Table S4: Parameters used for MultiXrank

### Supplementary Files

Supplementary File S1: Csv file containing the enrichment analysis results computed using the genes associated with physiological aging

Supplementary File S2: Excel file containing the gene nodes belonging to each of the 67 communities

Supplementary File S3: Excel file containing the gene nodes belonging to each cluster (i.e., the union of the genes belonging to the set of communities composing the cluster)

Supplementary File S4: Csv file containing the diseases belonging to each cluster

Supplementary File S5: Excel file containing the enrichment of clusters using lists of physiological aging genes

Supplementary File S6: Csv file containing the lists of genes differentially expressed in human blood, skin, brain, muscle and breast during aging, from the study of Irizar et al.

Supplementary File S7: Excel file containing the enrichment analysis results of the 67 communities, using GO Biological Processes and Cellular Components, and Reactome pathways functional annotations

Supplementary File S8: Excel file containing the enrichment analysis results of the 6 clusters, using GO Biological Processes and Cellular Components, and Reactome pathways functional annotations. The last 2 columns contain the seed nodes of the cluster annotated for the corresponding function and their corresponding diseases

Supplementary File S8bis: Excel file containing the enrichment analysis results of the 8 clusters obtained with an alternative cutoff of 0.5 in the dendrogram, using GO Biological Processes and Cellular Components, and Reactome pathways functional annotations.

Supplementary File S9: Excel file containing enrichment analysis results of the 6 clusters, using HPO phenotypes

### Supplementary Text

Supplementary Text S1: Comparison with alternative network exploration strategies, including classical (noniterative) Random Walk with Restart, Multilayer network partitioning, and computation of network distances with shortest paths.

## Notes

### Funding Statement

This work received support from:
The French National Research Agency (ANR-21-CE45-000101)
The Association Francaise contre les Myopathies (AFM)
The Excellence Initiative of Aix-Marseille University A*Midex (a French Investissements d’Avenir programme)
Institute MarMaRa AMX-19-IET-007
The European Union’s Horizon 2020 research and innovation programme under the EJP RD COFUND-EJP No 825575.
GPT-4 was used to improve syntax and readability.

### Author Declarations

The study used (or will use) ONLY openly available human data that were originally located at: – Orphanet: https://www.orpha.net/consor/cgi-bin/index.php – HuRI: http://www.interactome-atlas.org/download – APID: http://cicblade.dep.usal.es:8080/APID/init.action – Reactome: https://reactome.org/download-data – Human Protein Atlas: https://www.proteinatlas.org/about/download – CORUM: http://mips.helmholtz-muenchen.de/corum – hu.MAP: http://humap2.proteincomplexes.org/

### Summary of Updates

This revised version encompasses modifications and extensions of the text as well as new experiments comparing our network approaches to three alternative network exploration computational strategies. The novel experiments include: – The testing of an alternative cutoff threshold for cluster identification, which complements the different parameters tested for community sizes that were already in the initial version of the manuscript (Supplementary Figure S3). – The implementation and application of 3 alternative network exploration strategies, which include a classical (non-iterative) RWR, a multiplex-network partitioning algorithm (MolTi) and the computation of direct network distances with shortest paths. We added these novel implementations and comparisons as a supplementary text (Supplementary Text S1) containing the methods, results, and associated 3 supplementary figures (Supplementary Figures S4, S5, S6). Overall, the conclusion drawn from these additional experiments are that disease relationship signals are present in the networks. However, our iterative RWR algorithm reveals these signals more effectively than the other approaches. The supplementary material and associated conclusions have been added to the main text.

